# Adversity and patterns of health visiting in England: Evidence from linked administrative data and expert-by-experience workshops

**DOI:** 10.64898/2026.09.14.26362976

**Authors:** Louise Mc Grath-Lone, Katie Harron, Jane Barlow, Samantha Bennett, Sally Kendall, Jennifer Kirman, Alison Lamont, Mengyun Liu, Eirini-Christina Saloniki, Jenny Woodman

## Abstract

**Background:** Health visiting in England is a universal service designed to identify family need and offer more support where needed (proportionate universalism). We know little about how local areas deliver services for families facing adversity or their associated outcomes.

**Methods:** Using linked Community Services Data Set and Hospital Episode Statistics data (2018-20) for 57/152 areas, we (a) described health visiting service delivery for children whose mother had a previous hospital admission with mental health issues, alcohol/substance misuse or violence/abuse recorded (study definition of ‘adversity’), (b) identified common service delivery patterns using latent class analysis; and (c) tested whether patterns were associated with child and maternal outcomes, overall and in terms of inequalities for families facing adversity. We conducted four experts-by-experience workshops to aid interpretation.

**Results:** Three service delivery patterns for families facing adversity emerged, distinguished primarily by the intensity and age-distribution of contacts. In areas where health visiting was delivered with a high intensity across the whole 0-5 years age range (rather than targeted primarily at babies), children were more likely to meet expected levels of development at age 2-2½ years (OR_adj_: 0.79, 95% CI:0.63-0.99) and child inequalities in injury-related hospital admissions and some developmental domains were reduced.

**Conclusions:** We found that a pattern of health visiting aligned with proportionate universalism was associated with mitigating disadvantage for families facing adversity. With limited outcomes and forms of adversity captured in our study, we cannot say which service delivery pattern is ‘best’, but our findings evidence health visiting’s potential to reduce inequalities.

## Introduction

Health visiting is a universal preventive public health service for children aged 0-5 years in England through which skill-mix teams, led by specialist community public health nurses, deliver the Healthy Child Programme to families in their homes and in community and clinical settings.[1] Considerable variation in how health visiting services are delivered across local areas has been documented through national statistics and analyses of administrative data.[2–5] Understanding whether common patterns of service delivery exist despite local variation would support learning across contexts and the development of coherent national policy.

A core aim of health visiting is to reduce inequalities by addressing early disadvantage.[1] It is theorised to reduce the risk of adverse outcomes (such as childhood accidents and communication issues) through sustained, relationship-based contact, which supports parenting, identifies needs, and facilitates onward referral.[6] Health visiting is underpinned by the principle of proportionate universalism whereby every family receives a universal offer of five mandated contacts between 28 weeks of pregnancy and age 2½ years, with additional contacts provided to those with greater need.[5,7] In practice, additional contacts outnumber mandated contacts in most local areas.[3] Balancing the targeted additional support and the universal mandated offer has been identified as a key driver of local variation, particularly in the context of resource constraints, and is a research priority for the health visiting evidence base.[5,6]

As part of proportionate universalism, families facing adversity should receive more intensive health visiting support to mitigate adverse outcomes and reduce the social gradient. Consistent with this, recent evidence found children at greater risk of potentially avoidable hospital admissions by age 1 year (i.e., whose mothers had a history of adversity, specifically mental health issues, alcohol/substance misuse, or exposure to violence or abuse) were more likely to have received intensive health visiting in the first 30 days of life.[8,9] Evidence is lacking about patterns of service delivery across the full 0-5 years age range for families facing adversity, and whether different patterns can mitigate adverse outcomes.

This study uses linked Community Services Data Set (CSDS) and Hospital Episode Statistics (HES) data for 57 local authorities in England to: (a) describe how health visiting services are delivered up to age 5 years for families facing adversity; (b) identify whether common patterns of service delivery exist using latent class analysis (LCA); and (c) test whether patterns are associated with theoretically-relevant child and maternal outcomes, overall and in terms of inequalities for families facing adversity. This dual-estimand approach allows us to distinguish patterns of service delivery that benefit the population broadly from those that specifically mitigate the disadvantage faced by families facing adversity (i.e. that flatten the gradient between groups, rather than eliminate differences), thereby testing a premise that is closer to how proportionate universalism is intended to operate in practice.

## Methods

This analysis was part of a mixed-methods study[10] for which ethical approval was granted by University College London Institute of Education Research Ethics Committee (REC:1531).

### Data sources

CSDS includes details of health visiting contacts, including the location, duration and mode, results of developmental screening assessments, and child demographic information.[11] As the quality of CSDS data varies by financial quarter and local area,[12] we restricted our analyses to a sub-sample of 57 local authorities previously identified as having at least one quarter of data that was complete, relative to external reference data (Supplementary Table 1).[3] HES contains information about Accident & Emergency (A&E) attendances and hospital admissions to National Health Service hospitals, including admission date and type (e.g., emergency, elective), as well as clinical information captured using International Classification of Diseases 10^th^ Revision (ICD-10) diagnostic codes.[13] A common pseudonymised identifier enables linkage of children’s CSDS and HES records.[14]Mothers’ and babies’ HES records are also linkable.[15]

### Study populations

When describing and categorising health visiting services, we included children aged 0-5 years with a recorded contact between April 2018 and March 2020 in a local authority with complete CSDS data (Supplementary Figure 1). Adversity - the primary axis of inequality examined in this study - was derived from ICD-10 diagnosis codes related to mental health issues, alcohol/substance misuse and exposure to violence or abuse recorded in planned and unplanned hospital admissions in the 3 years prior to delivery (Supplementary Table 2), in line with previous studies.[8,9,16] When comparing outcomes, we included all live, singleton births recorded in HES between April 2018 and March 2019 that linked to CSDS (Supplementary Figure 2). We followed up each mother to their child’s 1^st^ birthday (longest period possible due to A&E data availability; Supplementary Table 3) and each child to their 5^th^ birthday (April 2023 to March 2024).

### Categorising health visiting service delivery for families facing adversity

For families facing adversity, we described characteristics of contacts overall, and by local authority to capture service delivery variation. This included the ratio of additional to mandated contacts, and the mode, location and duration of contacts. We used LCA to investigate whether common local authority-level patterns of service delivery could be identified. The assumption underlying LCA is that within a population there are distinct, homogenous subgroups (known as latent classes), and it is the mixture of these subgroups that accounts for the overall observed heterogeneity.[17] Latent class membership is conceptualised as a categorical variable that cannot be directly measured, but can be inferred from other measurable, categorical variables known as indicators. We created a set of 13 binary indicators that captured a local area’s relative position for all available characteristics of service delivery using distributional thresholds (e.g., 75^th^ centile; Supplementary Table 5). Using an established LCA framework which systematically tests whether an indicator should be included in the model based on changes to fit indices,[18] we selected the most parsimonious set. We then tested models with 2 to 10 latent classes selecting the final class number based on relative fit statistics, interpretability and class size (Supplementary Figure 3).[19] We assigned each local area to the class that best fit their service delivery pattern (i.e. highest probability of class membership) then compared service delivery across classes.

### Comparing child and maternal outcomes across health visiting service patterns

To explore whether different patterns of health visiting service delivery mitigate the poorer outcomes experienced by those facing adversity, we analysed data from HES/CSDS for the whole study population. We compared three outcomes across the identified service patterns: (a) frequent maternal A&E attendances before the child’s 1^st^ birthday, (b) unplanned injury-related hospital admission before age 5, and (c) not meeting the expected level of development at age 2-2½ years. These were the sub-set of outcomes from the original study protocol that were relevant to a “high-impact area” for health visiting,[20] theoretically amenable to health visiting,[6] and sufficiently common for statistical analysis using sub-national data (Supplementary Table 4). This protocol deviation was a response to the growing body of evidence related to outcome selection in administrative data when studying complex interventions.[9,21]

For A&E attendances, we selected a threshold used in previous studies of ‘frequent attenders’ (≥3 within 1 year).[22] Injury-related admissions were derived using established ICD-10 code lists[23,24] (Supplementary Table 6). Development at age 2-2½ years was based on domain-specific score cut-offs (Supplementary Table 7) for the 24-, 27- and 30-month Ages & Stages Questionnaire Third Edition (ASQ^®^-3).[25] The primary outcome was not meeting the overall expected level (i.e. any domain score below the cut-off)[26] and not meeting the level in each domain were included as secondary outcomes.

For each outcome, we calculated prevalence: overall, across service patterns and by adversity. We fit two logistic regression models; firstly, estimating the overall association between service patterns (exposure) and outcomes, and then exploring effect modification via interactions between service pattern and adversity, and adversity and deprivation. From the latter model, we estimated the adjusted risk difference between individuals exposed and unexposed to adversity within each service pattern (i.e. the average marginal effect) and compared pairwise differences. Both models used robust standard errors to account for clustering of individuals within local areas and adjusted for individual-level characteristics that varied in distribution across areas adopting different service patterns and were independently associated with the outcomes. This included: area-level deprivation (quintile of index of multiple deprivation), young motherhood (maternal age <25 years, a threshold used in previous studies),[8,9,16] first-time motherhood (parity of 0), ethnicity and, for child outcomes, sex. This analysis addressed a causal question (i.e. what is the risk difference in outcomes between service delivery patterns?) using an observational design; therefore, we interpreted adjusted associations cautiously accounting for the constraints this design places on causal inference.

### Experts-by-experience workshops

To contextualise and interpret the findings of the LCA, we conducted four experts-by-experience workshops with parents (N=27) with lived experience of the adversities relevant to the study (Table 1). The 90-minute workshops explored parent perspectives on whether, and why, distinguishing features of the three patterns of service delivery (number of contacts, child’s age at time of contacts, contact location) might affect service engagement and the impact of health visiting for families.

**Table 1:** Overview of experts-by-experience workshops.

| Type of adversity | Mothers who had experienced mental health difficulties | Fathers who had experienced, or supported their partner with, mental health difficulties | Mothers who had experienced alcohol / substance use in the home | Mothers who had experienced domestic violence and abuse |
| --- | --- | --- | --- | --- |
| **Format<sup>a</sup>** | In-person | Online | In-person | In-person |
| **Location** | London | National | North East England | South West England |
| **Number of participants** | 5 | 7 | 8 | 7 |
| **Ethnicity of participants** | Black British=2<br>Asian British=2<br>White American=1 | Not recorded<sup>b</sup> | White British=7<br>Asian British=1 | White British=6<br>Asian British=1 |
| **General approach** | To organise the workshops, we collaborated with national charities supporting parents with lived experience of the three forms of adversity relevant to this study. The workshop content and presentation materials were shared with the charity in advance to ensure appropriate content.<br>All workshops were led by a member of the research team and facilitated by the charity support staff. Participants were able to interrupt, challenge, skip some or all questions, and withdraw without explanation, and these opportunities were taken up in practice.<br>Discussions were not audio-recorded, but the researcher took hand-written notes during and immediately after each workshop. We did not inquire about the details of personal journeys and experiences, though sometimes these details were volunteered. Support staff were available during and after the workshop to help participants debrief and process the conversation as needed.<br>All participants who joined a workshop were gifted vouchers to thank them for their time and expertise. |  |  |  |

<sup>a</sup>We were guided by the charity partners when deciding on the workshop format. In-person workshops took place in locations regularly used by the charities which were familiar to participants. <sup>b</sup>Demographic details were collected via paper forms at in-person workshops but not captured for the online workshop.

## RESULTS

### Prevalence of adversity

Of all children aged 0-5 years in contact with health visiting services between April 2018 and March 2020 in the 57 included local authorities, 85.2% (533,585/626,480) had their birth recorded in HES and could be linked to a maternal delivery record (Supplementary Figure 1). Of these children, one in seven (13.8%) had an adversity-related maternal record in HES (Supplementary Table 8), namely mental health issues (13.3%), alcohol/substance misuse (1.5%) and violence or abuse (0.5%).

### Health visiting service delivery for children facing adversity

For children facing adversity (N=73,730), services delivered 2.4 additional contacts for every mandated contact (Table 2). Overall, 84.5% of contacts were delivered face-to-face with the majority recorded as home visits. Mandated contacts were more likely than additional contacts to be delivered face-to-face and at home. Additional contacts were most often delivered in the first 6 months of life (38.2%); however, one in five (22.1%) were delivered to children aged over 2½ years (i.e. after the final mandated contact). There was considerable local variation in service delivery for children facing adversity, including the duration of new birth visits and additional contacts, use of group sessions, and age-distribution of additional contacts (Table 2).

**Table 2:**
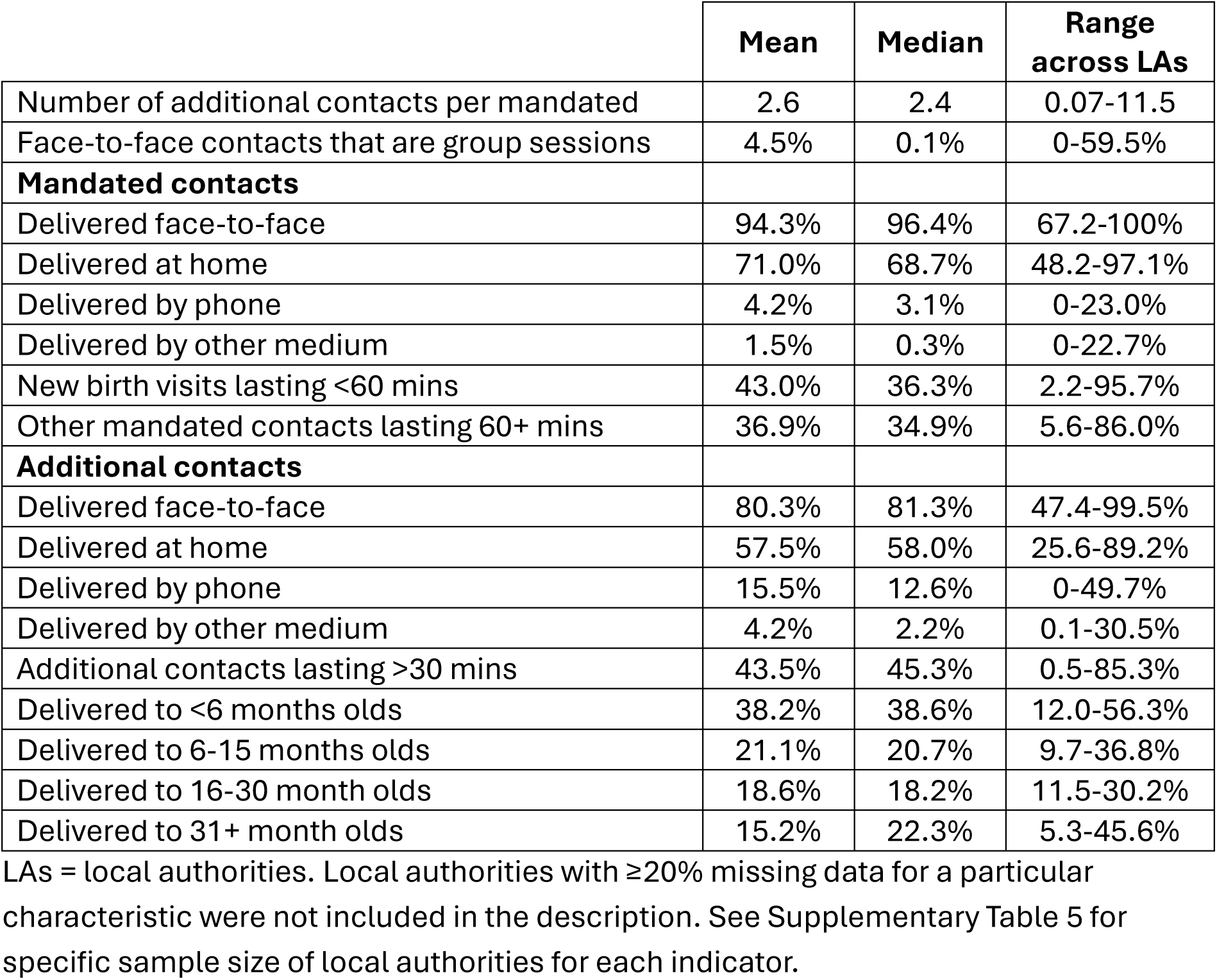
Local authority variation in characteristics of health visiting services delivered to families facing adversity between 1^st^ April 2018 to 31^st^ March 2020 (N=73,730)

|  | Mean | Median | Range across LAs |
| --- | --- | --- | --- |
| Number of additional contacts per mandated | 2.6 | 2.4 | 0.07-11.5 |
| Face-to-face contacts that are group sessions | 4.5% | 0.1% | 0-59.5% |
| **Mandated contacts** |  |  |  |
| Delivered face-to-face | 94.3% | 96.4% | 67.2-100% |
| Delivered at home | 71.0% | 68.7% | 48.2-97.1% |
| Delivered by phone | 4.2% | 3.1% | 0-23.0% |
| Delivered by other medium | 1.5% | 0.3% | 0-22.7% |
| New birth visits lasting <60 mins | 43.0% | 36.3% | 2.2-95.7% |
| Other mandated contacts lasting 60+ mins | 36.9% | 34.9% | 5.6-86.0% |
| **Additional contacts** |  |  |  |
| Delivered face-to-face | 80.3% | 81.3% | 47.4-99.5% |
| Delivered at home | 57.5% | 58.0% | 25.6-89.2% |
| Delivered by phone | 15.5% | 12.6% | 0-49.7% |
| Delivered by other medium | 4.2% | 2.2% | 0.1-30.5% |
| Additional contacts lasting >30 mins | 43.5% | 45.3% | 0.5-85.3% |
| Delivered to <6 months olds | 38.2% | 38.6% | 12.0-56.3% |
| Delivered to 6-15 months olds | 21.1% | 20.7% | 9.7-36.8% |
| Delivered to 16-30 month olds | 18.6% | 18.2% | 11.5-30.2% |
| Delivered to 31+ month olds | 15.2% | 22.3% | 5.3-45.6% |

### Patterns of health service delivery for children facing adversity

We identified three common service delivery patterns for children facing adversity (Table 3), primarily distinguished by differences in the ratio (intensity) and age-distribution (focus) of additional contacts. These were classified as: (a) lower intensity, 0-5 focus; (b) lower intensity, early focus; (c) higher intensity, 0-5 focus. Almost half of areas (46%) were classified as ‘lower intensity, early focus’. As well as the intensity and age-distribution of additional contacts, the three patterns differed in other aspects of service delivery. For example, ‘higher intensity, 0-5 focus’ areas delivered more face-to-face mandated contacts at home and more additional contacts by phone than other areas. ‘Lower intensity, 0-5 focus’ areas made more use of group sessions and other media (e.g., text messages) to deliver contacts. In both patterns with a 0-5 focus, over a quarter of additional contacts were delivered to children aged >2½ years (i.e. after the final mandated contact). ‘Lower intensity, early focus’ areas delivered 44% of additional contacts to children before the age of 6 months.

**Table 3:** Characteristics of common health visiting service patterns for families facing adversity that emerged from latent class analysis of the Community Services Data Set for 57 local authorities.

|  | **a) Lower intensity across full 0-5 years age range (N=20, 35%)** |  | **b) Lower intensity, focus on early intervention (N=26, 46%)** |  | **c) Higher intensity across full 0-5 years age range (N=11, 19%)** |  |
| --- | --- | --- | --- | --- | --- | --- |
| **Narrative description** | In these areas, additional contacts were more evenly distributed across the full 0-5 years age range and the ratio of additional to mandated contacts was lower, compared to other areas. Other aspects of service delivery that could be viewed as lower 'intensity' are: the greater use of group (rather than individual) contacts, greater use of other media for contacts (such as emails, texts, etc. rather than phone or in-person) and higher proportion of new birth visits lasting <60 minutes. |  | In these areas, additional contacts were concentrated in the 0-6 months age range and the ratio of additional to mandated contacts was lower, compared to other areas. Other aspects of service delivery that could be viewed as lower 'intensity' are: higher proportion of additional contacts lasting <30 minutes. |  | In these areas, additional contacts were more evenly distributed across the full 0-5 years age range and the ratio of additional to mandated contacts was higher, compared to other areas. Other aspects of service delivery that could be viewed as higher 'intensity' are: the very low use of group (rather than individual) contacts, very low use of other media for contacts (such as emails, texts, etc. rather than phone or in-person), higher proportion of mandated visits delivered at home and lasting 60+ minutes and higher proportion of additional visits delivered at home. |  |
| **Overall** | **Mean** | **IQR** | **Mean** | **IQR** | **Mean** | **IQR** |
| Number of additional contacts per mandated | 2.4 | 1.31-2.44 | 2.4 | 1.95-3.10 | 3.4 | 2.10-3.99 |
| Face-to-face contacts that are group sessions | 10.1% | 0.03-16.0% | 2.9% | 0-0.08% | 0.2% | 0.0-0.0% |
| **Mandated contacts** | **Mean** | **IQR** | **Mean** | **IQR** | **Mean** | **IQR** |

|  | a) Lower intensity across full 0-5 years age range (N=20, 35%) |  | b) Lower intensity, focus on early intervention (N=26, 46%) |  | c) Higher intensity across full 0-5 years age range (N=11, 19%) |  |
| --- | --- | --- | --- | --- | --- | --- |
| Delivered face-to-face | 93.2% | 91.8-97.2% | 95.1% | 94.2-98.1% | 94.6% | 93.4-96.8% |
| Delivered at home | 65.8% | 58.5-72.8% | 66.6% | 61.2-72.4% | 90.7% | 85.2-96.5% |
| Delivered by phone | 4.1% | 1.7-7.2% | 4.1% | 1.6-5.3% | 4.7% | 3.0-6.1% |
| Delivered by other medium | 2.7% | 0.03-1.98% | 0.8% | 0.03-0.99% | 0.6% | 0.1-1.1% |
| New birth visits lasting <60 mins | 31.3% | 10.2-41.9% | 47.6% | 20.1-70.7% | 56.0% | 28.8-93.0% |
| Other mandated contacts lasting 60+ mins | 39.0% | 28.0-46.8% | 29.5% | 18.1-36.5% | 52.3% | 35.7-58.9% |
| **Additional contacts** | **Mean** | **IQR** | **Mean** | **IQR** | **Mean** | **IQR** |
| Delivered face-to-face | 75.3% | 68.8-82.3% | 88.1% | 78.1-93.6% | 74.4% | 50.5-88.7% |
| Delivered at home | 58.8% | 50.1-67.9% | 47.7% | 38.9-57.6% | 76.3% | 71.2-79.4% |
| Delivered by phone | 17.4% | 12.6-22.9% | 12.5% | 4.82-19.2% | 22.2% | 9.0-47.4% |
| Delivered by other medium | 7.3% | 2.3-13.0% | 2.5% | 0.3-2.7% | 3.0% | 2.1-3.2% |
| Additional contacts lasting >30 mins | 49.4% | 44.2-60.3% | 37.5% | 21.1-48.5% | 47.8% | 33.4-57.1% |
| Delivered to <6 months olds | 33.7% | 31.5-37.7% | 44.0% | 41.4-45.9% | 32.5% | 27.6-36.9% |
| Delivered to 6-15 months olds | 18.7% | 17.2-21.7% | 23.4% | 20.2-24.8% | 19.6% | 18.3-20.7% |
| Delivered to 16-30 month olds | 20.1% | 17.0-22.1% | 17.0% | 14.9-19.5% | 19.8% | 16.6-22.6% |
| Delivered to 31+ month olds | 27.5% | 24.3-30.6% | 15.5% | 13.1-19.2% | 28.1% | 22.8-33.1% |

IQR = Interquartile range (25<sup>th</sup> to 75<sup>th</sup> centile). Shading highlights defining features of the latent class ('service pattern') relative to other classes.

### Insights from experts-by-experience workshops

Both mothers and fathers in workshops covering mental health viewed the role of health visiting as valuable for identifying difficulties and offering meaningful support. Mothers in particular identified frequent additional contacts in the home, especially in the perinatal period, to be easier to engage with than clinic contacts which were reported as stressful. Both mothers and fathers emphasised the benefit of continuity with one health visiting professional for feeling understood.

In contrast, participants in the workshop for families experiencing addiction did not see the value of additional health visiting contacts as they reported that these families would likely already have other service involvement. Furthermore, home visits were considered likely to be stressful, as parents would feel under scrutiny for signs of alcohol/substance use, whether or not they were using. Participants suggested parents experiencing addiction in the home may seek to minimise health visiting contact as much as possible without raising suspicion.

Home visits were also considered to be potential sources of stress for mothers experiencing domestic violence or abuse as they felt under surveillance at home. Home visits also risked the health visiting professional meeting the abuser, who mothers felt may be able to win their confidence. Clinic visits were viewed favourably as a legitimate opportunity for mothers to leave the house which may otherwise be difficult. While visits in the perinatal period were considered important for mothers experiencing domestic violence or abuse, participants highlighted the changing pressures of parenting as the baby becomes a toddler, which they viewed as a period where health visiting could be useful for disclosure and/or support.

### Maternal and child outcomes

Of the relevant study populations (Supplementary Figure 2), 2.6% mothers had ≥3 A&E attendances before their baby’s 1^st^ birthday, 5.5% of children had an unplanned injury-related hospital admission by age 5 years, and 11.9% did not meet the expected level of development at age 2-2½ years. Poorer outcomes were more common amongst younger mothers and those who lived in the most deprived areas, were not first-time mothers, and were facing adversity (Supplementary Table 9).

Prevalence of outcomes varied across service patterns, with the highest unadjusted rates of frequent maternal A&E visits and child injury admissions in the ‘higher intensity, 0-5 focus’ areas, and the highest rates of not meeting the expected levels of development in the ‘lower intensity, early focus’ areas (Table 4). After adjusting for co-variates, differences across service patterns for frequent maternal A&E visits and child injury admissions were attenuated and no longer statistically significant (Table 4). There was a 4.5 percentage point crude difference in frequent A&E attendances between mothers exposed and unexposed to adversity (6.5% vs 2.0%), but the adjusted risk difference across service patterns were not statistically significant (Figure 1, Supplementary Table 10). There was a 2.6 percentage point gap in injury-related admissions between children exposed and unexposed to adversity overall (7.7% vs 5.1%) which was significantly smaller in ‘higher intensity, 0-5 focus’ areas compared to others (Figure 1, Supplementary Table 10).

**Figure 1:**
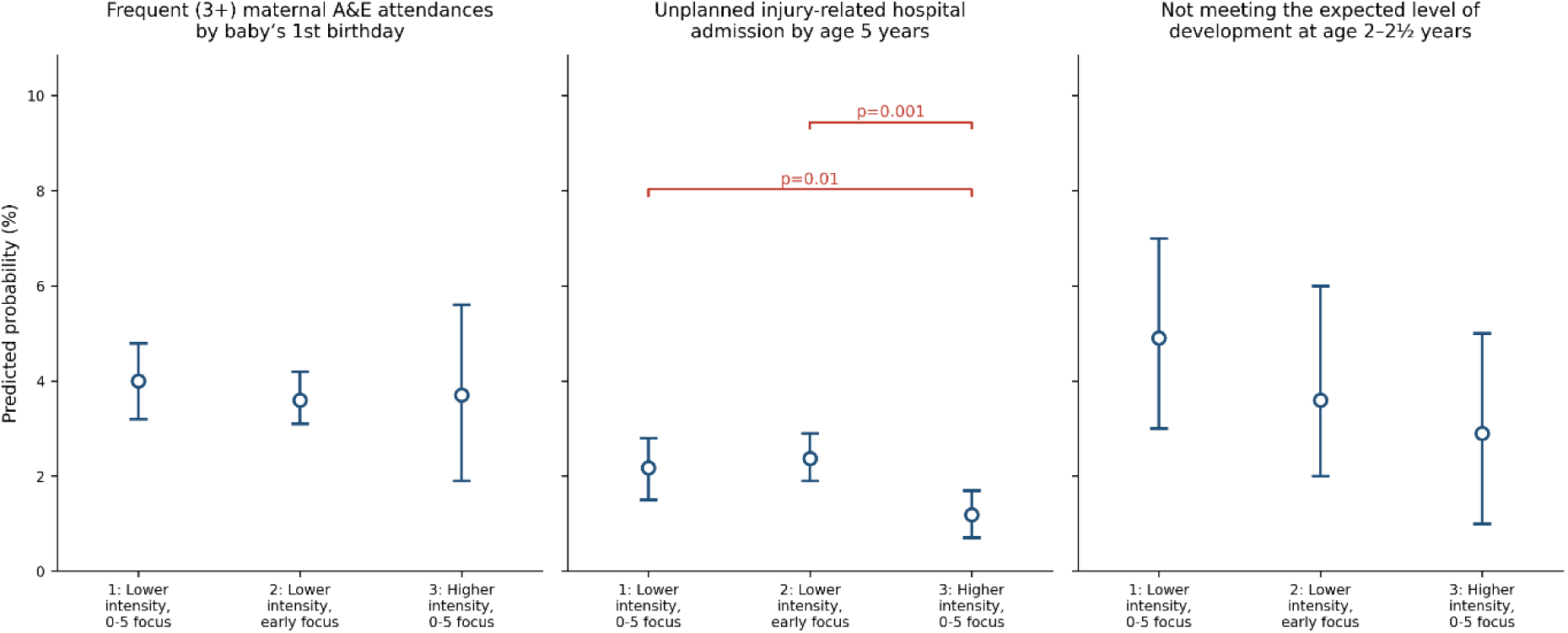
Inequalities in maternal and child outcomes for families facing adversity, by service pattern: predicted margins with 95% confidence intervals. A&E = Accident & Emergency. Brackets highlight pairwise differences in outcomes between service patterns with a p-value <0.05 - see Supplementary Table 10 for further details.

**Table 4:** Maternal and child outcomes by health visiting service pattern.

| **Frequent (3+) maternal A&E attendances by baby's 1<sup>st</sup> birthday (N=202,840)** |  |  |  |  |  |  |  |
| --- | --- | --- | --- | --- | --- | --- | --- |
| **Service pattern** | **Prevalence** | **OR** | **95% CI** | **p-value** | **OR<sub>adj</sub><sup>a</sup>** | **95% CI** | **p-value** |
| A: Lower intensity, 0-5 focus | 2.4% | (ref) |  |  | (ref) |  |  |
| B: Lower intensity, early focus | 2.4% | 1.02 | 0.96-1.08 | 0.56 | 0.99 | 0.78-1.27 | 0.95 |
| C: Higher intensity, 0-5 focus | 3.7% | 1.58 | 1.46-1.71 | **<0.001** | 1.12 | 0.71-1.77 | 0.63 |
| **Unplanned injury-related hospital admission by age 5 years (N=196,050)** |  |  |  |  |  |  |  |
| **Service pattern** | **%** | **OR** | **95% CI** | **p-value** | **OR<sub>adj</sub><sup>b</sup>** | **95% CI** | **p-value** |
| A: Lower intensity, 0-5 focus | 5.3% | (ref) |  |  | (ref) |  |  |
| B: Lower intensity, early focus | 5.3% | 1.01 | 0.96-1.05 | 0.82 | 0.99 | 0.85-1.14 | 0.86 |
| C: Higher intensity, 0-5 focus | 6.3% | 1.21 | 1.14-1.28 | **<0.001** | 1.12 | 0.94-1.34 | 0.22 |
| **Not meeting the expected level of development at age 2-2 ½ years (N=41,690)** |  |  |  |  |  |  |  |
| **Service pattern** | **%** | **OR** | **95% CI** | **p-value** | **OR<sub>adj</sub><sup>b</sup>** | **95% CI** | **p-value** |
| A: Lower intensity, 0-5 focus | 10.9% | (ref) |  |  | (ref) |  |  |
| B: Lower intensity, early focus | 13.3% | 1.25 | 1.17-1.34 | **<0.001** | 1.11 | 0.82-1.51 | 0.49 |
| C: Higher intensity, 0-5 focus | 11.1% | 1.02 | 0.94-1.11 | 0.62 | 0.79 | 0.63-0.99 | **0.04** |

A&E = Accident & Emergency; OR = odds ratio; CI = confidence interval. Bold indicates p-value <0.05. <sup>a</sup>Adjusted for ethnicity, deprivation, being a young mother (<25 years), being a first-time mother (parity = 0), and being exposed to adversity. <sup>b</sup>Adjusted for sex, ethnicity, deprivation, being a young mother (<25 years), being a first-time mother (parity = 0), and being exposed to adversity.

Children in ‘higher intensity, 0-5 focus’ areas had 21% lower adjusted odds of not meeting the expected level of development than those in areas with a ‘lower intensity, 0-5 focus’ approach (Table 4). There was a 4.4 percentage point crude gap in not meeting the expected level of development between children exposed and unexposed to adversity (15.7% vs 11.3%), but differences across service patterns were not statistically significant. Secondary analyses focusing on the specific domains comprising the composite development measure showed mitigation effects of the ‘higher intensity, 0-5 focus’ in personal-social, problem-solving, and gross motor domains, which had smaller gaps compared to other areas (Supplementary Table 11).

## DISCUSSION

Our study is the first to describe how local services in England deliver health visiting across the full 0-5 years age range to families facing adversity. We found that additional contacts played a substantial role for families facing adversity (median ratio of 2.4 for each mandated contact) and included high proportions delivered face-to-face (81.3%), at home (58.0%), and lasting more than 30 minutes (45.3%). The experts-by-experience workshops suggested that health visiting was seen as an acceptable and helpful intervention by mothers who had experienced mental health issues. However, the service was seen as a more problematic intervention by mothers who had lived with alcohol or substance misuse or domestic violence and abuse, with constraints on how far mothers could have an honest conversation with the health visiting professional, for different reasons. Honest conversation and a trusting relationship are one of the key mechanisms by which health visiting is theorised to improve child and family health. The mothers even suggested that for some women health visiting may be harmful; for example, if the abuser aligned themselves with the health visiting professional.

Our findings also show that patterns in local health visiting service delivery can be identified. Local areas appear to be adopting recognisable, common approaches to service delivery, even in the absence of an explicit framework for doing so. This raises the question of whether service patterns emerge organically as an accumulated response to local need and operational pressures, or are adopted strategically with commissioners and providers considering which best fits the needs of their population and the outcomes they are seeking to influence. Although the delivery patterns identified in this study are not definitive, they indicate how health visiting services may enact proportionate universalism in different ways and may be useful to guide planning and research.

Our finding that ‘higher intensity, 0-5 focus’ service delivery (a pattern consistent with health visiting’s theoretical mechanism) is associated with mitigating disadvantage in some outcomes for families facing adversity provides empirical evidence of the service’s potential to reduce inequalities. The inequality in unplanned hospital admissions for injuries between children in families facing adversity was approximately half that observed in the areas with ‘lower intensity’ service patterns. ‘Higher intensity, 0-5 focus’ areas also had significantly lower overall adjusted odds of not meeting the expected level of development at age 2–2½ years as assessed by ASQ®-3 and there were signs of narrowing gaps in specific aspects of child development in ‘higher intensity, 0-5 focus’ LAs, specifically personal-social, problem-solving, and gross motor domains. We found no association between service pattern and frequent maternal A&E attendances; however, this must be interpreted in terms of the non-random allocation of service patterns. For example, the ‘higher intensity, 0-5 focus’ service pattern was more likely to be adopted in higher-need areas, so the attenuation of unadjusted associations with A&E attendances has two potential interpretations that the study design cannot distinguish. It may be that the service pattern has no effect, or it may be that it successfully offset the elevated underlying risk in the areas that adopted this pattern. The mitigation finding for injury admissions strengthens the latter interpretation.

A strength of this study is the use of LCA to identify varying patterns of service delivery for families facing adversity which provide commissioners and practitioners with a description more closely aligned to how services are designed, operated and experienced. These insights may help commissioners and practitioners reflect on how to deliver services to specific groups they aim to engage and support. This approach is also helpful because it provides a framework for understanding and informing national policy. Comparisons to learn across contexts are otherwise hindered by the overwhelming amount of variation that suggests each local area is doing something uniquely different (or, as has previously been described by stakeholders, that there are “152 ways of [delivering health visiting services]”).[4] A further strength is the dual-estimand framing of our outcome analyses which examined both overall odds across the whole population and the adversity-related gaps. The distinction between health visiting’s broad and targeted influences is useful because services may be commissioned on the basis of population-level benefit, mitigation of inequalities, or both. Evaluating a complex intervention such as health visiting using administrative data requires careful outcome selection: indicators with weak theoretical links risk producing uninterpretable or spurious findings.[21] Restricting our analyses to outcomes aligned with high-impact areas for health visiting[20] and with clear conceptual grounding in its logic model[6] strengthens the empirical robustness and policy relevance of our findings.

There are several methodological limitations. Firstly, our operationalisation of adversity (identified from ICD-10 diagnosis codes related to mental health, alcohol/substance misuse and violence in mothers’ hospital admission records in the three years prior to delivery), although consistent with previous administrative data studies[8,9,16] captures only a subset of the population burden. For example, adversity arising after a child was born or which was known to primary care or other services (but not hospitals) is excluded. As such, the 13.8% of children in our sample classified as facing adversity is likely an underestimate of the true vulnerable population. It is unclear what effect this underestimation may have on the direction or magnitude of bias: a broader operationalisation may strengthen observed mitigation effects by reducing bias toward the null, or attenuate effects if the population facing forms of adversity that are identifiable from hospital records have the greatest capacity to benefit from intensive health visiting support. A second limitation is that the three service patterns we identified through LCA are a statistical simplification of what is, in reality, a continuous landscape of variation in delivery. Local areas were assigned to the pattern that best fitted their configuration of binary indicators based on distributional cut-offs of continuous variables that are recorded in administrative data. There is variation within each service pattern and they should be understood as heuristic analytical categories that help to describe and compare complex services, rather than a definitive or fixed typology. Finally, service pattern is not randomly allocated: LA provision is a response to perceived need, which means comparing across patterns conflates the pattern itself with unobserved area-level characteristics that influenced its adoption. Adjustment for individual-level covariates partly addresses compositional differences, and the mitigation analysis is more robust than the overall comparison because the difference between groups is estimated within each area. Nonetheless, residual confounding cannot be excluded.

Existing evidence shows families facing adversity receive more health visiting support in the first year of life than their peers, predominantly through additional contacts: 10% of children facing adversity have 10 or more additional contacts by age 1.[8] Our analysis extends this picture to the full 0-5 age range and shows that additional contacts continue to play a substantial role beyond infancy. A sizeable, but varying, proportion of additional contacts are delivered after the final mandated contact at age 2½ years; this is a period of particular policy relevance with the government’s current focus on improving school readiness by the end of Reception year in primary school (age 4-5 years).[27] The only previous large-scale empirical test of health visiting outcomes in England examined the effect of intensive health visiting in the first 30 days of life on avoidable hospital admissions during infancy,[9] but the positive association between hospital admissions and having received more intensive health visiting was difficult to interpret: results may have been explained by residual confounding, selection bias, or effects of health visiting to increase family health-seeking behaviour in hospitals. In contrast, our findings are broadly coherent with the theoretical mechanism through which health visiting is thought to operate.[6] Specifically, the ‘higher-intensity, 0-5 focus’ service pattern, which was associated with mitigation of adverse outcomes for children facing adversity, was distinguished by more face-to-face mandated contacts at home, longer contacts, and additional contacts distributed across the full 0-5 age range. These are features that align with sustained, relationship-based contact that supports parenting, identifies emerging needs, and facilitates onward referral. However, in our experts-by-experience workshops, mothers warned that this way of working with families may not be possible when there are addictions or domestic violence and abuse. This was also the least common pattern of delivery across the included areas.

Further research is needed to understand how decisions about health visiting commissioning, service design, delivery and monitoring are made in practice, including the role of administrative data at local and national levels. Strengthening CSDS data quality would allow our analyses to be extended to more local areas improving the generalisability of these findings, and further linkage to education and other health data would help to determine whether the service patterns we observed are associated with differences in children’s life trajectories. This would enable us to build a more complete understanding of how, in services based on proportionate universalism, variation emerges and translates into outcomes for the children and families, including those who most need support.

## Data availability statement

No data are available. The data underlying this article cannot be shared publicly due to the terms of the data-sharing agreement with NHS England. Data can be obtained by submitting a data request through the NHS England Data Access Request Service.

