## Supplemental Material for "Adversity and patterns of health visiting in England: Evidence from linked administrative data and expert-by-experience workshops"

### Supplemental Table 1: Characteristics of local authorities included and not included in the analysis dataset, N (%)

| Characteristic | 149 local authorities in England[a] | 57 local authorities included in analysis | 93 local authorities not included in analysis | p-value[b] |
|---|---|---|---|---|
| **Region** | | | | 0.79 |
| East Midlands | 9 (6.0) | 4 (7.0) | 5 (5.4) | |
| East of England | 12 (8.1) | 4 (7.0) | 8 (8.7) | |
| London | 32 (21.5) | 8 (14.0) | 24 (26.1) | |
| North East | 12 (8.1) | 6 (10.5) | 6 (6.5) | |
| North West | 23 (15.4) | 8 (14.0) | 15 (16.3) | |
| South East | 18 (12.1) | 8 (14.0) | 10 (10.9) | |
| South West | 14 (9.4) | 7 (7.5) | 7 (12.3) | |
| West Midlands | 14 (9.4) | 6 (10.5) | 8 (8.7) | |
| Yorkshire and the Humber | 15 (10.1) | 6 (10.5) | 9 (9.8) | |
| **Urban / rural status** [c] | | | | 0.79 |
| Predominantly rural | 20 (13.4) | 9 (15.8) | 11 (12.0) | |
| Predominantly urban | 108 (72.5) | 40 (70.2) | 68 (73.9) | |
| Urban with significant rural | 21 (14.1) | 8 (14.0) | 13 (14.1) | |
| **Income Deprivation Affecting Children Index (IDACI) quintiles** | | | | 0.92 |
| Lowest quintile | 30 (20.1) | 12 (21.1) | 18 (19.6) | |
| 2nd quintile | 30 (20.1) | 10 (17.5) | 20 (21.7) | |
| 3rd quintile | 30 (20.1) | 11 (19.3) | 19 (20.7) | |
| 4th quintile | 30 (20.1) | 11 (19.3) | 19 (20.7) | |
| Highest quintile | 29 (19.5) | 13 (22.8) | 16 (17.4) | |

[a]The total number of local authorities was 149 as City of London is combined with Hackney and Isles of Scilly is combined with Cornwall. [b]When testing the difference between the local authorities included vs. not included in the analysis, p-values were calculated using Fisher's exact test for categorical values. [c]Urban/rural status based on 2011 3-fold classification. [d]Deprivation based on 2019 quintiles summarised from individual score ranks.

### Supplemental Table 2: International Classification of Diseases 10th Revision (ICD-10) diagnostic codes for adversity

| Adversity | ICD-10 codes | ICD-10 Description |
|---|---|---|
| Maternal mental health | F320 | Mild depressive episode |
| | F321 | Moderate depressive episode |
| | F322 | Severe depression without psychotic symptoms |
| | F328 | Other depressive episodes |
| | F329 | Depressive episode, unspecified |
| | F330 | Recurrent depressive disorder, current episode mild |
| | F331 | Recurrent depressive disorder, current episode moderate |
| | F332 | Recurrent depressive disorder, current episode severe without psychotic symptoms |
| | F333 | Recurrent depressive disorder, current episode severe with psychotic symptoms |
| | F338 | Other recurrent depressive disorders |
| | F339 | Recurrent depressive disorder, unspecified |
| | F340 | Cyclothymia |
| | F341 | Dysthymia |
| | F348 | Other persistent mood (affective) disorders |
| | F349 | Persistent mood (affective) disorder, unspecified |
| | F380 | Other single mood (affective) disorders |
| | F381 | Other recurrent mood (affective) disorders |
| | F388 | Other specified mood (affective) disorders |
| | F39 | Unspecified mood (affective) disorder |
| | F40 | Phobic anxiety disorders |
| | F41 | Other anxiety disorders |
| | F42 | Obsessive-compulsive disorder |
| | F44 | Dissociative (conversation) disorders |
| | F45 | Somatoform disorders |
| | F48 | Other neurotic disorders |
| | F54 | Psychological and behavioural factors associated with disorders or diseases classified elsewhere |
| | F94.0 | Elective mutism |
| | F43 | Reaction to severe stress, and adjustment disorders |
| | F62 | Enduring personality changes, not attributable to brain damage and disease |
| | F94.1 | Reactive attachment disorder of childhood |
| | F94.2 | Disinhibited attachment disorder of childhood |
| | F50 | Eating disorders |
| | F51 | Non-organic sleep disorders |
| | F55 | Abuse of non-dependence-producing substances |
| | F93 | Emotional disorders with onset specific to childhood |
| | F94 | Disorders of social functioning with onset specific to childhood and adolescence |

| Adversity | ICD-10 codes | ICD-10 Description |
|---|---|---|
| | F98 | Other behavioural and emotional disorders with onset usually occurring in childhood and adolescence |
| | F63 | Habit and impulse disorders |
| | F90 | Disturbance of activity and attention |
| | F91 | Conduct disorders |
| | F92 | Mixed disorders of conduct and emotions |
| | F30 | Manic episode |
| | F31 | Bipolar affective disorder |
| | F323 | Severe depression with psychotic symptoms |
| | F20 | Schizophrenia |
| | F21 | Schizotypal disorder |
| | F22 | Persistent delusional disorders |
| | F23 | Acute and transient psychotic disorders |
| | F24 | Induced delusional disorder |
| | F25 | Schizoaffective disorders |
| | F28 | Other nonorganic psychotic disorders |
| | F29 | Unspecified nonorganic psychosis |
| | F60 | Specific Personality Disorders |
| | F61 | Mixed and other personality disorders |
| | F68 | Other disorders of adult personality and behaviour |
| | F69 | Unspecified disorder of adult personality and behaviour |
| | F53 | Mental & behavioural disorders associated with the puerperium |
| | F10 | Mental and behavioural disorders due to use of alcohol |
| Alcohol / substance misuse | F11-16, F18-19 | Mental and behavioural disorders due to psychoactive substance use |
| | R78.0 | Finding of alcohol in blood |
| | R78.1- R78.5 | Findings of drugs and other substances, not normally found in blood |
| | T36-T50 | Poisoning by drugs, medicaments and biological substances |
| | X45 | Accidental poisoning by and exposure to alcohol |
| | X40-X44 X46-X49 | Accidental poisoning by and exposure to noxious substances |
| | X69 | Intentional self-poisoning by and exposure to other and unspecified chemicals and noxious substances |
| | Y10-Y14, Y16-Y19 | Poisoning, undetermined intent |
| | Y15 | Poisoning by and exposure to alcohol, undetermined intent |
| | Y90 | Evidence of alcohol involvement determined by blood alcohol level |
| | Y91 | Evidence of alcohol involvement determined by level of intoxication |
| | Z50.2 | Alcohol rehabilitation |
| | Z50.3 | Drug rehabilitation |

| Adversity | ICD-10 codes | ICD-10 Description |
|---|---|---|
| | Z71.4 | Alcohol abuse counselling and surveillance |
| | Z71.5 | Drug abuse counselling and surveillance |
| | Z72.1 | Problems related to lifestyle, alcohol use |
| | Z72.2 | Problems related to lifestyle, drug use |
| | G40.5 | Special epileptic syndromes |
| | Z04.0 | Examination for Blood-alcohol and blood-drug test |
| | O35.4 | Maternal care for (suspected) damage to foetus from alcohol |
| | E24.4 | Alcohol-induced pseudo-Cushing syndrome |
| | G31.2 | Degeneration of nervous system due to alcohol |
| | G62.1 | Alcoholic polyneuropathy |
| | G72.1 | Alcoholic polyneuropathy |
| | I42.6 | Alcoholic cardiomyopathy |
| | K29.2 | Alcoholic gastritis |
| | K70 | Alcoholic liver disease |
| | K85.2 | Alcohol-induced acute pancreatitis |
| | K86.0 | Alcohol-induced chronic pancreatitis |
| | T51 | Toxic effect of alcohol |
| Violence (including domestic violence & self-harm) | T74 | Maltreatment syndrome |
| | T73 | Effects of other deprivation (extreme neglect) |
| | Y06, Y07 | Perpetrator of neglect and other maltreatment syndromes |
| | Y04, Y05 | Assault by bodily force and sexual assault |
| | X85- Y03, Y08, Y09 | Other type of assault |
| | Y20-Y34 | Events of undetermined intent |
| | Z04.5 | Examination and observation following other inflicted injury |
| | Z04.8 | Examination and observation for other reasons: request for expert evidence |
| | Z63.0 | Problems in relationship with spouse or partner |
| | X60-68 | Intentional self-poisoning |
| | X70-X84 | Intentional self-harm |
| | Y87.0 | Sequelae of intentional self-harm |
| | Z91.5 | Personal history of self-harm |

Adversity was defined as an admission that included one or more the above codes in any diagnostic position in the three years prior to delivery. For alcohol/substance misuse and violence, only emergency (unplanned) admissions were considered.

### Supplemental Table 3: Availability of data sources at time of analysis in relation to cohort selection

| | | Calendar year | | | | | | |
|---|---|---|---|---|---|---|---|---|
| | | **2018/19** | **2019/20** | **2020/21** | **2021/22** | **2022/23** | **2023/24** | **2024/25** |
| **Year of birth** | **2018/19** | *0* | *1* | *2* | *3* | *4* | *5* | *6* |
| | **2019/20** | | *0* | *1* | *2* | *3* | *4* | *5* |
| | **2020/21** | | | *0* | *1* | *2* | *3* | *4* |
| **Data source** | **CSDS** | Yes | Yes | Yes | Yes | No | No | No |
| | **HES APC** | Yes | Yes | Yes | Yes | Yes | Yes | No |
| | **HES A&E** | Yes | Yes | No | No | No | No | No |

CSDS = Community Services Data Set; HES = Hospital Episodes Statistics; APC = Admitted Patient Care; A&E = Accident & Emergency; ECDS = Emergency Care Dataset. N/A = Not available at the time of analysis (June 2025). Italicised numbers indicate age in years for relevant birth cohort. Orange shading indicates issues with data availability in 2020/21, namely that the HES A&E data collection was replaced with ECDS; however, a different pseudonymised person-level identifier was used which prevented longitudinal records of A&E visits being examined beyond 31st March 2020. Green box highlights the 2018/19 cohort selected for outcome analyses.

### Supplemental Table 4: Assessment of candidate maternal and child outcomes

| Outcome indicator | Theoretical relevance | Sufficiently powered | Decision |
|---|---|---|---|
| 1. Unplanned hospital admission (child) | No: too crude to be considered an indicator of 'Reduction in avoidable and unnecessary hospital use' | Unlikely to be an issue: not a rare outcome | **Exclude**: not theoretically relevant |
| 2. Injury/ingestion-related admission (child) | Yes: a (sharp end) indicator of 'Reduced child accidents and injuries' | Unlikely to be an issue: not a rare outcome | **Include:** theoretically relevant, analysis likely to be sufficiently powered |
| 3. Maltreatment-related admission (child) | Yes: a (sharp end) indicator of 'Reduced child maltreatment' | Likely to be an issue: relatively rare outcome | **Exclude**: analysis likely to be underpowered |
| 4. Mortality (child) | No: not included in health visiting logic model | No: very rare outcome | **Exclude**: not theoretically relevant, analysis likely to be underpowered |
| 5. Expected level of child development | Yes: an indicator of 'Improved child language and communication' | Unlikely to be an issue: not a rare outcome | **Include:** theoretically relevant, analysis likely to be sufficiently powered |
| 6. Repeated A&E attendances within 1 year (child) | No: too crude to be considered an indicator of 'Reduction in avoidable and unnecessary hospital use' | Unlikely to be an issue: not a rare outcome | **Exclude**: not theoretically relevant |
| 7. Unplanned hospital admission (mother) | No: too crude to be considered an indicator of 'Reduction in avoidable and unnecessary hospital use' | Unlikely to be an issue: not a rare outcome | **Exclude**: not theoretically relevant |
| 8. ACEs-related admission (mother) | No: too crude to be considered an indicator of 'Reduced Parental alcohol and drug use' | Likely to be an issue: relatively rare outcome | **Exclude**: not theoretically relevant, analysis likely to be underpowered |
| 9. Repeated A&E attendances within 1 year (mother) | Yes: an indicator of 'Reduction in avoidable and unnecessary hospital use' | Unlikely to be an issue: not a rare outcome | **Include:** theoretically relevant, analysis likely to be sufficiently powered |

Nine outcome indicators that can be created using Community Services Data Set or Hospital Episodes Statistics were considered for inclusion in the analysis. Outcome selection was guided by two criteria: a) theorised as amenable to intervention by health visiting services with reference to the published logic model for health visiting and b) common enough to be likely to provide sufficient statistical power to identify differences between sub-groups, where they exist, in the sub-national data with reference to published prevalence figures.

### Supplemental Table 5: Indicators of health visiting service delivery derived from Community Service Data Set and considered for inclusion in the latent class analysis

| **Relative indicator description** | **Binary cut-off** | **Threshold cut-off** | **Local authorities with >20% contacts have missing data n (%)** | **Included in analysis?[a]** |
|---|---|---|---|---|
| Higher ratio of additional to mandated contacts | ≥3 | ≥75th centile | 0 | Yes |
| Greater use of face-to-face groups sessions | ≥10% | ≥90th centile | 8 (16.3%) | Yes |
| More face-to-face contacts at medical or health setting | ≥25% | ≥75th centile | 8 (16.3%) | No |
| More additional contacts delivered to proxy (parent, carer) | ≥15% | ≥75th centile | 0 | Yes |
| **Mandated contacts** | | | | |
| Lower use of face-to-face mandated contacts | <90% | ≤10th centile | 4 (7.5%) | Yes |
| More mandated contacts at home | ≥80% | ≥75th centile | 9 (18.8%) | Yes |
| More new birth visits lasting ≥60 minutes | ≥66% | ≥75th centile | 6 (11.8%) | Yes |
| More long mandated contacts (≥60 minutes) | ≥50% | ≥75th centile | 6 (11.8%) | Yes |
| **Additional contacts** | | | | |
| Lower use of face-to-face additional contacts | <75% | ≤25th centile | 13 (29.5%) | Yes |
| More face-to-face additional contacts at home | ≥66% | ≥75th centile | 8 (16.3%) | Yes |
| More long additional contacts (≥30 minutes) | ≥66% | ≥75th centile | 4 (7.5%) | Yes |
| Higher use of additional contacts at <6 months olds | ≥40% | ≥75th centile | 0 | Yes |
| Higher use of additional contacts at ≥31 month olds | ≥25% | ≤25th centile | 0 | Yes |

Continuous service characteristics were dichotomised at local authority level using distributional thresholds. This data-driven approach to indicator creation captures the relative position of local authorities against the national distribution. [a]When deciding whether an indicator should be included in the latent class model, we used the Dean & Rafferty framework which systematically tests whether the addition of an indicator improves model fit indices.

### Supplemental Table 6: International Classification of Diseases 10th Revision (ICD-10) diagnostic codes for injury-related admission

| ICD-10 codes | ICD-10 Description |
|---|---|
| S00-S09 | Injuries to the head (includes open wounds, fractures, crushing and dislocation) |
| S10-S19 | Injuries to the neck |
| S20-S29 | Injuries to the thorax |
| S30-S39 | Injuries to the abdomen, lower back, lumbar spine and pelvis |
| S40-S49 | Injuries to the shoulder and upper arm |
| S50-S59 | Injuries to the elbow and forearm |
| S60-S69 | Injuries to the wrist and hand |
| S70-S79 | Injuries to the hip and thigh |
| S80-S89 | Injuries to the knee and lower leg |
| S90-S99 | Injuries to the ankle and foot |
| T00-T07 | Injuries involving multiple body regions |
| T08-T14 | Injuries to unspecified part of trunk, limb or body region |
| T15-T19 | Effects of foreign body entering through natural orifice |
| T20-T32 | Burns and corrosions |
| T33-T35 | Frostbite |
| T36-T50 | Poisoning by drugs, medicaments and biological substances |
| T51-T65 | Toxic effects of substances chiefly non-medicinal as to source (sting, alcohol, solvents, etc.) |
| T66-T78 | Other and unspecified effects of external causes (effects of radiation, heat and light, hypothermia, electric shock, asphyxiation, food deprivation) |
| X40-X49 | Accidental poisoning by and exposure to noxious substances |
| T79 | Certain early complications of trauma |
| T90-T98 | Sequelae of injuries, of poisoning and of other consequences of external causes |
| V01-V99 | Transport accidents |
| W00-W19 | Falls |
| W20-W49 | Exposure to inanimate mechanical forces |
| W50-W64 | Exposure to animate mechanical forces |
| W65-W74 | Accidental drowning and submersion |
| W75-W84 | Other accidental threats to breathing |
| W85-W99 | Exposure to electric current, radiation, and extreme ambient air temperature |
| X00-X09 | Exposure to smoke, fire and flames |
| X10-X19 | Contact with heat and hot substances |
| X58-X59 | Accidental exposure to other and unspecified factors |
| Y10-19 | Events of undetermined intent (poisonings) |
| Y20-Y34 | Events of undetermined intent (hanging, drowning, firearm discharge…) |
| Y35-Y36 | Legal intervention and operations of war |
| Y85 | Sequelae of transport accidents |
| Y86, Y87.2, Y89.9 | Sequelae of other accidents/events of undetermined intent/unspecified external causes |
| Y89.0, Y89.1 | Sequelae of legal intervention and war operations |

Injury-related admission was defined as a planned or unplanned admission that included one or more the above codes in any diagnostic position.

### Supplemental Table 7: Version- and domain-specific cut-off scores for Ages & Stages Questionnaire 3rd Edition (ASQ-3) relevant to 2-2½ year olds

| | Version | | |
|---|---|---|---|
| **Domain** | **24-month** | **27-month** | **30-month** |
| **Communication** | 25.17 | 24.02 | 33.30 |
| **Gross motor** | 38.07 | 28.01 | 36.14 |
| **Fine motor** | 35.16 | 18.42 | 19.25 |
| **Problem-solving** | 29.78 | 27.62 | 27.08 |
| **Personal-social** | 31.54 | 25.31 | 32.01 |

As published in supporting documentation for the administration of Ages & Stages Questionnaire 3rd Edition (ASQ-3) versions relevant to 2-2½ year olds.

### Supplemental Table 8: Characteristics of children receiving health visiting services between 1st April 2018 to 31st March 2020 (N=533,585)

| **Sex**[a] | **n** | **%** | **First language** | **n** | **%** |
|---|---|---|---|---|---|
| Boys | 278,550 | 52.2% | English | 167,045 | 31.3% |
| Girls (or unknown) | 255,030 | 47.8% | Other | 16,620 | 3.1% |
| | | | Unknown | 349,915 | 65.6% |
| **Ethnicity**[b] | | | | | |
| White | 372,355 | 69.8% | **Type of adversity**[c] | | |
| Asian | 38,500 | 7.2% | Maternal mental health | 71,065 | 13.3% |
| Black | 14,250 | 2.7% | Alcohol/substance misuse | 8,065 | 1.5% |
| Mixed or Other | 51,395 | 9.6% | Violence | 2,710 | 0.5% |
| Unknown | 57,085 | 10.7% | Any of the above | 73,730 | 13.8% |

[a]Children of unknown sex were combined with 'girls' to comply with the data provider's statistical disclosure requirement to not report findings based on cell size <10. [b]Ethnicity was categorised based on the standard 16+1 ethnicity codes recorded in administrative health datasets. [c]Adversity was derived from International Classification of Diseases 10th Revision (ICD-10) diagnostic codes` related to mental health, substance misuse and violence captured in planned and unplanned hospital admission records in the 3 years prior to delivery (see Supplemental Table 2).

### Supplemental Table 9: Prevalence of primary outcomes by maternal and child characteristics

| | Frequent (3+) maternal A&E attendances by baby's 1st birthday (N=202,840) | | Unplanned injury-related hospital admission by age 5 years (N=196,050) | | Not meeting the expected level of development at age 2-2 ½years[a] (N=41,690) | |
|---|---|---|---|---|---|---|
| **Sex** | **%** | **p-value** | **%** | **p-value** | **%** | **p-value** |
| Boys | n/a | | 6.0% | | 16.1% | |
| Girls (or unknown)[b] | n/a | n/a | 4.9% | **<0.001** | 7.5% | **<0.001** |
| **Ethnic category[c]** | | | | | | |
| White | 2.8% | | 5.7% | | 11.5% | |
| Asian | 2.3% | | 4.5% | | 16.3% | |
| Black | 3.5% | | 4.3% | | 17.0% | |
| Mixed or Other ethnicity | 3.6% | **<0.001** | 5.2% | **<0.001** | 11.4% | **<0.001** |
| **Deprivation quintile[d]** | | | | | | |
| 1st (least deprived) | 1.4% | | 4.7% | | 7.8% | |
| 2nd quintile | 1.8% | | 5.3% | | 9.8% | |
| 3rd quintile | 2.2% | | 5.4% | | 11.2% | |
| 4th quintile | 2.9% | | 5.4% | | 12.6% | |
| 5th (most deprived) | 4.2% | **<0.001** | 6.2% | **<0.001** | 15.7% | **<0.001** |
| **Young mother?[e]** | | | | | | |
| No | 2.0% | | 5.0% | | 11.1% | |
| Yes | 5.5% | **<0.001** | 7.6% | **<0.001** | 16.3% | **<0.001** |
| **First time mother?[f]** | | | | | | |
| No | 2.7% | | 5.7% | | 11.8% | |
| Yes | 2.4% | **<0.001** | 5.2% | **<0.001** | 12.0% | |
| **Exposed to adversity?[g]** | | | | | | |
| No | 2.0% | | 5.1% | | 11.3% | |
| Yes | 6.5% | **<0.001** | 7.7% | **<0.001** | 15.7% | **<0.001** |
| **Type of adversity[g]** | | | | | | |
| Maternal mental health | 6.5% | **<0.001** | 7.7% | **<0.001** | 15.8% | **<0.001** |
| Alcohol/substance misuse | 14.5% | **<0.001** | 9.8% | **<0.001** | 19.1% | **<0.001** |
| Violence | 14.1% | **<0.001** | 10.0% | **<0.001** | 20.9% | **<0.001** |

A&E = Accident & Emergency. Bold highlights p-value <0.05 based on $chi^2$ test. [a]Expected levels of development assessed using 24-, 27- and 30-month Ages & Stages Questionnaire 3rd Edition (ASQ-3). [b]Children of unknown sex were combined with 'girls' to comply with the data provider's statistical disclosure requirement to not report findings based on cell size <10. [c]Ethnicity was categorised based on 16+1 ethnicity code recorded in administrative health datasets. 'Unknown' ethnicity category is not reported. [d]Based on area-level Index of Multiple Deprivation at the time of birth. [e]Young motherhood defined as maternal age <25 years. [f]First-time motherhood defined as parity of 0. [g]Adversity was derived from International Classification of Diseases 10th Revision (ICD-10) diagnostic codes` related to mental health, substance misuse and violence captured in planned and unplanned hospital admission records in the 3 years prior to delivery (Supplemental Table 2).

### Supplemental Table 10: Pairwise comparisons of adjusted risk differences (adversity-exposed vs unexposed mothers/children) between health visiting service patterns (primary outcomes)

| **Frequent (3+) maternal A&E attendances by baby's 1st birthday (N=202,840)** | **Difference in PP** | **95% CI** | **p-value** |
|---|---|---|---|
| A vs B: Lower intensity, early focus versus Lower intensity, 0-5 focus | -0.3 | -1.3 to 0.6 | 0.47 |
| C vs A: Higher intensity, 0-5 focus versus Lower intensity, early focus | -0.3 | -2.2 to 1.7 | 0.80 |
| C vs B: Higher intensity, 0-5 focus versus Lower intensity, 0-5 focus | 0.1 | -1.8 to 2.0 | 0.91 |
| **Unplanned injury-related hospital admission by age 5 years (N=196,050)** | **Difference in PP** | **95% CI** | **p-value** |
| A vs B: Lower intensity, early focus versus Lower intensity, 0-5 focus | 0.2 | -0.06 to 1.0 | 0.62 |
| C vs A: Higher intensity, 0-5 focus versus Lower intensity, early focus | -1.0 | -1.7 to -0.02 | **0.01** |
| C vs B: Higher intensity, 0-5 focus versus Lower intensity, 0-5 focus | -1.2 | -1.9 to -0.05 | **0.001** |
| **Not meeting the expected level of development at age 2-2 ½years[a] (N=41,690)** | **Difference in PP** | **95% CI** | **p-value** |
| A vs B: Lower intensity, early focus versus Lower intensity, 0-5 focus | -1.3 | -4.1 to 1.5 | 0.36 |
| C vs A: Higher intensity, 0-5 focus versus Lower intensity, early focus | -2.0 | -4.7 to 0.7 | 0.15 |
| C vs B: Higher intensity, 0-5 focus versus Lower intensity, 0-5 focus | -0.6 | -3.2 to 2.0 | 0.62 |

A&E = Accident & Emergency; PP = percentage points; CI = confidence intervals. Bold highlights p-value <0.05. [a]Expected levels of development assessed using 24-, 27- and 30-month Ages & Stages Questionnaire 3rd Edition (ASQ-3).

**Supplemental Table 11: Pairwise comparisons of adjusted risk differences (adversity-exposed vs unexposed children) between health visiting service patterns (secondary child development outcomes, N=41,690)**

| **Communication domain** | **Difference in PP** | **95% CI** | **p-value** |
|---|---|---|---|
| A vs B: Lower intensity, early focus versus Lower intensity, 0-5 focus | -1.5 | -4.1 to 0.7 | 0.19 |
| C vs A: Higher intensity, 0-5 focus versus Lower intensity, 0-5 focus | -1.5 | -3.9 to 1.0 | 0.25 |
| C vs B: Higher intensity, 0-5 focus versus Lower intensity, early focus | -0.01 | -2.0 to 2.1 | 0.99 |
| **Problem-solving domain** | **Difference in PP** | **95% CI** | **p-value** |
| A vs B: Lower intensity, early focus versus Lower intensity, 0-5 focus | -0.9 | -2.0 to 0.6 | 0.233 |
| C vs A: Higher intensity, 0-5 focus versus Lower intensity, 0-5 focus | -2.0 | -5.3 to -0.08 | **0.04** |
| C vs B: Higher intensity, 0-5 focus versus Lower intensity, early focus | -1.0 | -2.0 to 0.4 | 0.21 |
| **Personal social domain** | **Difference in PP** | **95% CI** | **p-value** |
| A vs B: Lower intensity, early focus versus Lower intensity, 0-5 focus | -1.1 | -2.9 to 0.5 | 0.18 |
| C vs A: Higher intensity, 0-5 focus versus Lower intensity, 0-5 focus | -3.0 | -4.3 to -0.8 | **0.004** |
| C vs B: Higher intensity, 0-5 focus versus Lower intensity, early focus | -1.4 | -3.1 to -0.03 | **0.04** |
| **Fine motor domain** | **Difference in PP** | **95% CI** | **p-value** |
| A vs B: Lower intensity, early focus versus Lower intensity, 0-5 focus | -0.06 | -2.1 to 1.4 | 0.934 |
| C vs A: Higher intensity, 0-5 focus versus Lower intensity, 0-5 focus | -0.8 | -2.3 to 0.6 | 0.255 |
| C vs B: Higher intensity, 0-5 focus versus Lower intensity, early focus | -0.7 | -2.2 to 1.0 | 0.414 |
| **Gross motor domain** | **Difference in PP** | **95% CI** | **p-value** |
| A vs B: Lower intensity, early focus versus Lower intensity, 0-5 focus | -1.2 | -3.4 to 0.6 | 0.18 |
| C vs A: Higher intensity, 0-5 focus versus Lower intensity, 0-5 focus | -2.0 | -4.1 to -0.3 | **0.02** |
| C vs B: Higher intensity, 0-5 focus versus Lower intensity, early focus | -1.0 | -3.3 to 0.5 | 0.18 |

The secondary outcomes explored were the five domains of development that comprise the Ages & Stages Questionnaire 3rd Edition (ASQ-3). PP = percentage points; CI = confidence interval. Bold highlights p-value <0.05.

### Supplemental Figure 1: Service description study cohort flow chart

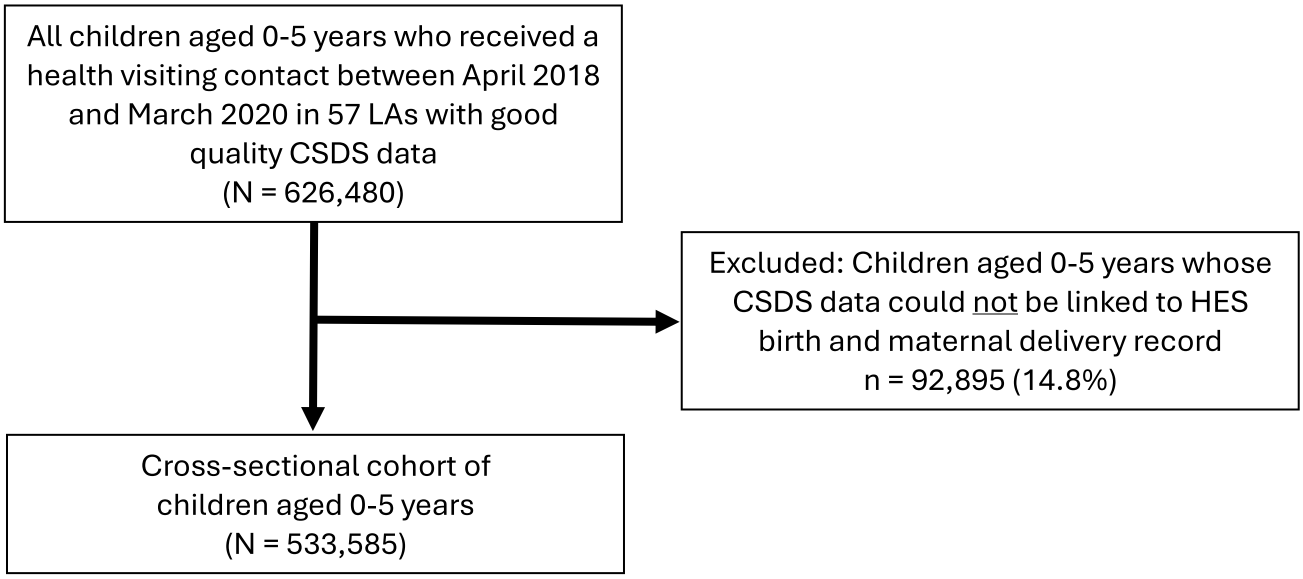

Due to data provider requirements related to statistical disclosure checks, all values are reported rounded to the closest 5. Therefore, numbers may not sum accurately across stages of the flowchart.

### Supplemental Figure 2: Outcomes study cohort flow chart

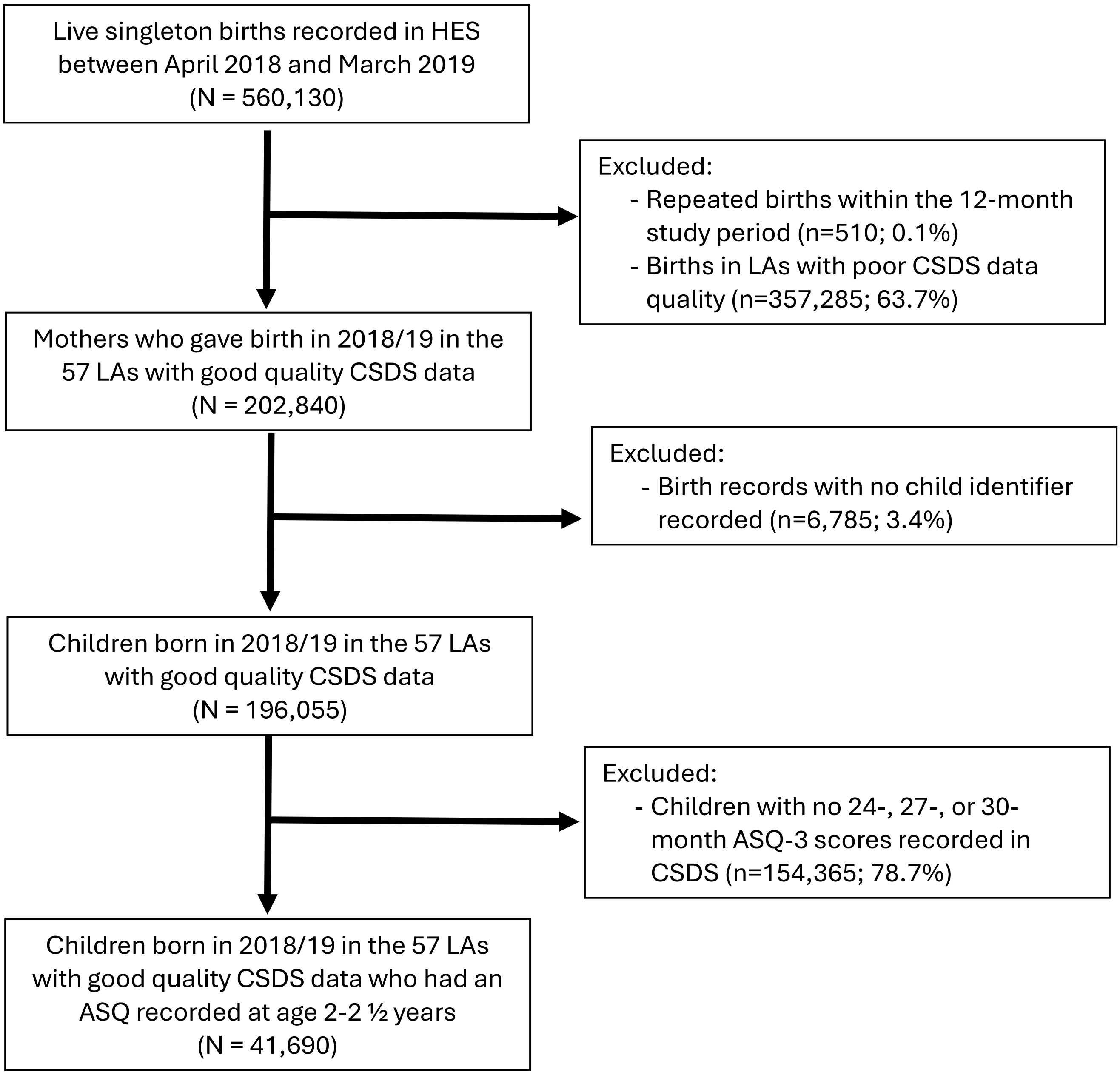

Due to data provider requirements related to statistical disclosure checks, all values are reported rounded to the closest 5. Therefore, numbers may not sum accurately across stages of the flowchart.

### Supplemental Figure 3: Fit indices for latent class model with 2 to10 classes

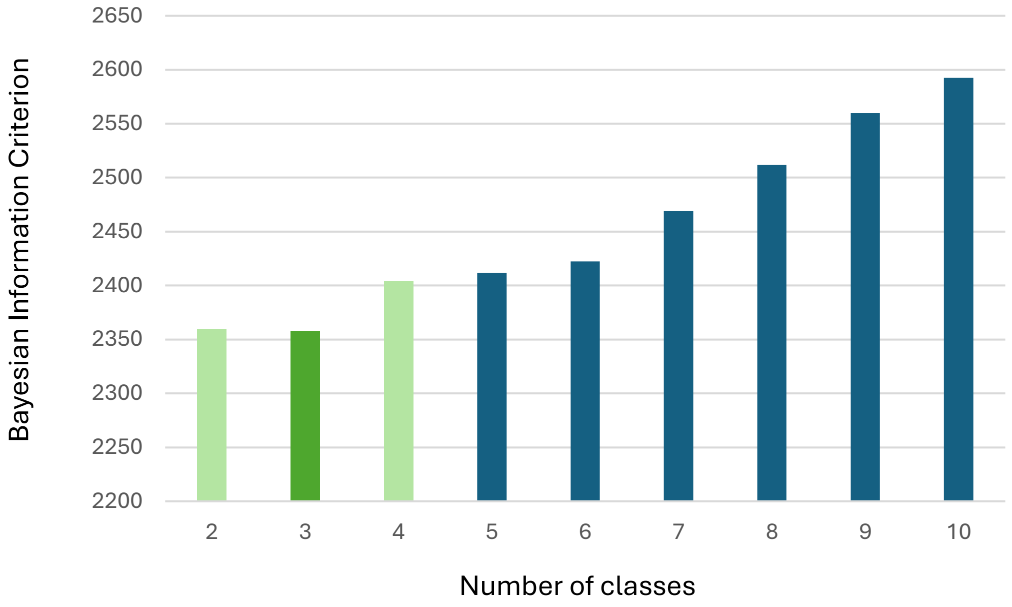

When choosing the optimum class-solution for a latent class model, lower values of the Bayesian Information Criterion (BIC) indicate better fit. Green shading highlights the class solutions (2 to 4) that were considered in terms of their interpretability and size when choosing the final class-solution.
